# “*Being proactive*”: A qualitative study of South Asian Australians perspectives of cardiovascular disease and genomic testing

**DOI:** 10.1101/2025.06.26.25330306

**Authors:** Emily DeBortoli, Vaishnavi Nathan, Heena Akbar, Aideen McInerney-Leo, Deborah Gilroy, Anjali Henders, Sonia Shah, Tatiane Yanes, the South Asian Genes and Health in Australia (SAGHA) research team

## Abstract

**Objectives:** Explore the health beliefs of South Asian Australians related to cardiovascular disease (CVD) and genomic testing for CVD

**Design, setting, participants:** Qualitative study using focus groups conducted between April to August 2023. Participants included individuals who self-identified as South Asian, aged 18 years or older, and resided in Queensland, Australia at the time of recruitment.

**Main outcome:** South Asian Australian health beliefs related to CVD and genomic testing for CVD

**Results:** A total of 60 individuals consented to participate (*n=*9 focus groups, and *n=*1 interview). All participants had lived experience with CVD, which shaped their related health beliefs. For some participants, these experiences contributed to significant disease related worry and reinforced fatalistic beliefs. Participants were aware of the multifactorial nature of CVD, however, recognised that cultural norms, age and gender affected engagement in regular health checks. Most participants understood the role of genomics in CVD and held positive attitudes towards genomic testing. However, several participants raised concerns about the effectiveness of genomic testing for individuals from diverse backgrounds, as well as challenges related to insurance coverage and data security.

**Conclusions:** Future CVD prevention strategies should consider South Asian Australians’ health beliefs, and the unique factors involved in incorporating genomic information to refine risk.

**Summary Box:** *The known:* South Asians have a two-fold greater risk of cardiovascular disease (CVD) than White-Europeans. In Australia, South Asians represent the largest, non-European immigrant population and a thus a key demographic for CVD risk management.

*The new:* South Asian Australian participants recognised that CVD risk is modifiable, however engagement in preventive behaviours varied and was shaped by cultural and demographic factors. Attitudes toward genomic testing were positive, with recognition that testing could refine risk assessments, although distinct considerations influence decision-making.

*The implications:* Future Australian CVD interventions should account for South Asians health beliefs about CVD and genomic testing.

## Introduction

Cardiovascular disease (CVD) is the leading cause of mortality in Australia, accounting for 24% of deaths annually (1). CVD risk is based on an interplay of genetic, environmental and lifestyle factors (2). South Asians (ancestral origins from India, Bangladesh, Sri Lanka, Pakistan, etc.) experience a higher risk of CVD compared to other ancestry groups (3). Specifically, South Asians have a two-fold greater risk of CVD compared to White-Europeans, with elevated risk remaining after adjusting for traditional risk factors (3). This population is also at an increased risk from a younger age and experience higher mortality rates (3-5). In Australia, South Asians constitute the largest, non-European migrant population (6), and thus, are a key demographic for CVD risk management.

Identifying individuals at high-risk of CVD allows for tailored risk management (e.g. cholesterol-lowering treatments and lifestyle adjustments) (7). In Australia, the AusCVDRisk calculator (8) is used in primary care to identify high-risk individuals (8, 9). However, the AusCVDRisk tool may not accurately capture CVD risk for individuals of non-European ancestry, including South Asians, as it does not account for population risk differences (10). Polygenic risk scores (PRS) (i.e., combined impact of multiple, low-risk genetic variants) have been shown to improve CVD risk estimates when integrated into risk tools (9). However, PRS has reduced accuracy for South Asian populations comparative to those of European ancestry due to under-representation in genomic databases (11). Consequently, non-European individuals at high-risk of CVD who would benefit from risk-modifying interventions may be overlooked due to inaccurate risk estimates. Improving the representation of diverse communities in genomic databases is widely recognised as a priority area (12, 13).

Little is known about the health beliefs and attitudes of South Asians towards CVD and genomic testing. Available studies indicate that South Asians perceive themselves at higher risk of CVD due to cultural influences, lifestyle factors, and limited access to healthcare (14, 15). Despite the high proportion of South Asians living in Australia (i.e., 4.1% of the Australian population are born in South Asia) (6), no genomic studies have focused on this population to date. However, South Asians health beliefs about genomic testing have been explored in other contexts internationally, including pharmacogenomics and cancer (16, 17). The South Asian Genes and Health in Australia (SAGHA) study, established in 2023, aims to improve the participation of South Asian Australians in genomic and CVD research (18). Key aspect of SAGHA included extensive community consultations, a consumer advisory group, and establishing community relationships. This study aimed to explore South Asian Australians experience with, and attitudes towards CVD and genomic testing.

## Methods

### Study design and setting

This study employed a qualitative design, grounded in phenomenology methodology and pragmatic paradigm. These methodology decisions enabled flexible method selection that aligned with the aims and facilitated an in-depth exploration of individuals’ lived experiences and perspectives. Focus groups were selected to facilitate dynamic discussions, generate data on collective views, and provide insight into the reasoning and experiences that shape these perspectives (19).

### Participant eligibility and recruitment

Eligible participants self-identified as South Asian aged 18 years or older, and resided in Queensland, Australia at the time of recruitment. Though countries such as Nepal and Bhutan are geographically considered to be part of South Asia, their genetic ancestry is distinct with East Asian admixture. Previous studies have shown East Asians to have lower CVD risk compared to those of European ancestry (20). For these reasons, this study focused on South Asians who self-reported to be of Indian, Bangladeshi, Sri Lankan and/or Pakistani ancestry. Individuals were eligible to participate regardless of English proficiency, with funding available for interpreters.

An online expression of interest (EOI) form was disseminated via the research team, members of the community advisory group, and community leaders. The EOI detailed the study purpose, research team, and the voluntary nature of participation. Recruitment was further facilitated by study team members speaking at community engagement events and connecting with community leaders, closed-group advertising (i.e. University of Queensland newsletter, LinkedIn and community social media groups), and word-of-mouth. A purposeful sampling approach applied to ensure a broad cross-section of the community based on gender, age, and South Asian ethnic groups. Snowball sampling was also applied, with some participants volunteering to be community liaisons and connecting the researcher team with interested members of their community. Individuals provided consent and completed a demographic questionnaire prior to participating in a focus group. All participations received a $30 gift card as compensation of their time.

### Data collection

Focus groups were conducted between April and August 2023, either virtually (i.e., Zoom) or in-person at various locations in Brisbane, Australia, including community venues and places of worship. Given the importance of group interactions and shared experiences, community-specific focus groups were conducted. Data collection continued until representation from all ancestral groups were collected. One participant was independently interviewed due to scheduling conflicts. All focus groups were conducted by South Asian authors V.N. and S.S. and facilitated by community advocates authors R.N. and H.A. Guided by discussions with community leaders, gender specific focus groups options were provided to facilitate open discussions that were tailored to cultural gender specific context.

Focus groups commenced with introductions, followed by a brief presentation on the purpose of the study, and later a second presentation on genomics and CVD risk, that included a jar models to depict PRS (21) (Supplementary Material 1). A semi-structured interview guide was used to promote discussion on topics including CVD, related health beliefs, and perceptions towards genomic testing (Supplementary Materials 2). Focus groups and the individual interview were audio-recorded, transcribed verbatim, and de-identified. All study information was securely stored on the University of Queensland servers and only accessible by the study team.

### Data Analysis and Synthesis

An inductive thematic analysis was undertaken using the methodology described by Braun and Clarke (22), with NVivo version 11 (23) used to sort common themes. Transcripts were coded by authors: H.A. (Pacific public health researcher of Fijian/South Asian ancestry), V.N. (research genetic counsellor of Indian ancestry), E.D. (research genetic counsellor of White-European ancestry) and T.Y. (genetic counsellor and researcher of Brazilian ancestry). Initial codes were grouped into preliminary themes, after which, sub-themes were developed, merged, removed or separated as supported by the data and group discussions, including thematic mapping.

### Ethics approval

This study received ethics approval from the University of Queensland Human Research Ethics Committee (UQ2022/HE001388). Findings are reported in accordance with the Standards for Reporting Qualitative Research.

## Results

### Participant characteristics

Of the 78 individuals who submitted an EOI, 60 (77%) consented to participate (*n=*9 focus groups, and *n=*1 interview). Among non-participants who submitted an EOI, one individual was deemed ineligible due to residing interstate, two withdrew after providing consent, and 13 were unavailable or did not reply to invitations to attend a focus group. Additionally, two individuals required an interpreter, which the study team was not made aware of and thus could not be accommodated for the focus group. Among the 60 participants, most had Indian ancestry (*n=*43; 72%), identified as female (*n=*39; 65%) and were between 30 to 49 years (*n=*35; 58%) (Table 1). Focus group discussions were categorised into three key themes: i) common thread of CVD, ii) empowering healthcare, and iii) genomics is predictive, not definitive. Representative quotes are displayed in Table 2, where pseudonyms have been applied, and are visually presented in Figure 1.

**Table 1:** Participant Demographic Characteristics.

| Demographic Characteristics | n | % |
| --- | --- | --- |
| *Biological sex* |  |  |
| Female | 39 | 65 |
| Male | 21 | 35 |
| *Ancestry* |  |  |
| India | 43 | 72 |
| Bangladesh | 7 | 12 |
| Sri Lanka | 5 | 8 |
| Pakistan | 5 | 8 |
| *Self-reported ethnicity\** |  |  |
| Bengali | 3 | 5 |
| Gujarati | 1 | 2 |
| Marathi | 1 | 2 |
| Indo-Fijian | 15 | 25 |
| Kannadigas | 1 | 2 |
| Malayali | 2 | 3 |
| Punjabi | 12 | 20 |
| Sinhalese | 2 | 3 |
| Tamil | 9 | 15 |
| Telugu | 2 | 3 |
| Not reported^ | 15 | 25 |
| *Age* |  |  |
| 18-29 | 7 | 12 |
| 30-39 | 23 | 38 |
| 40-49 | 12 | 20 |
| 50-59 | 6 | 10 |
| 60+ | 12 | 20 |

\*values sum more than 100% as individuals reported having more than one ethnicity; ^not reported at time of EOI

**Table 2:**
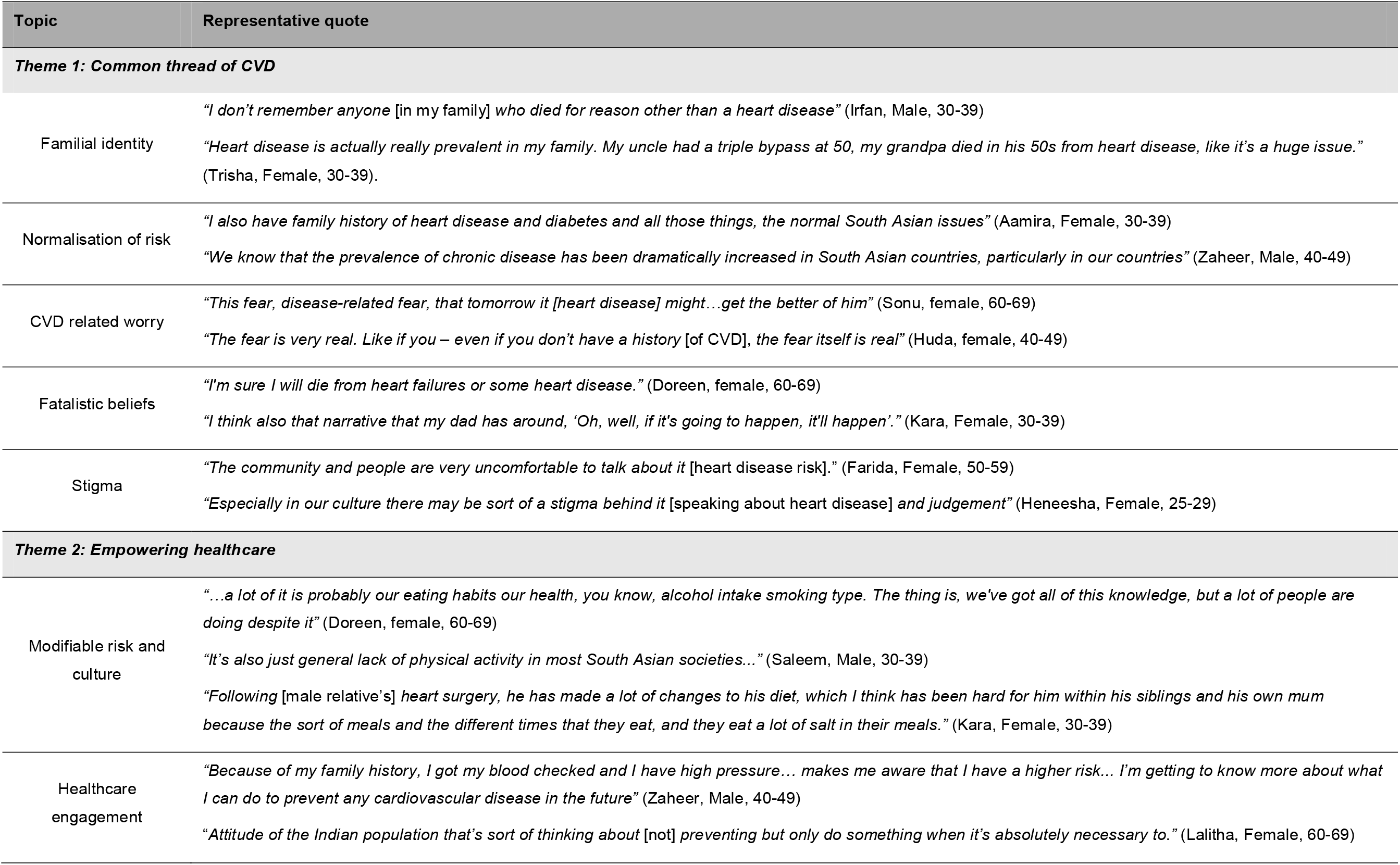
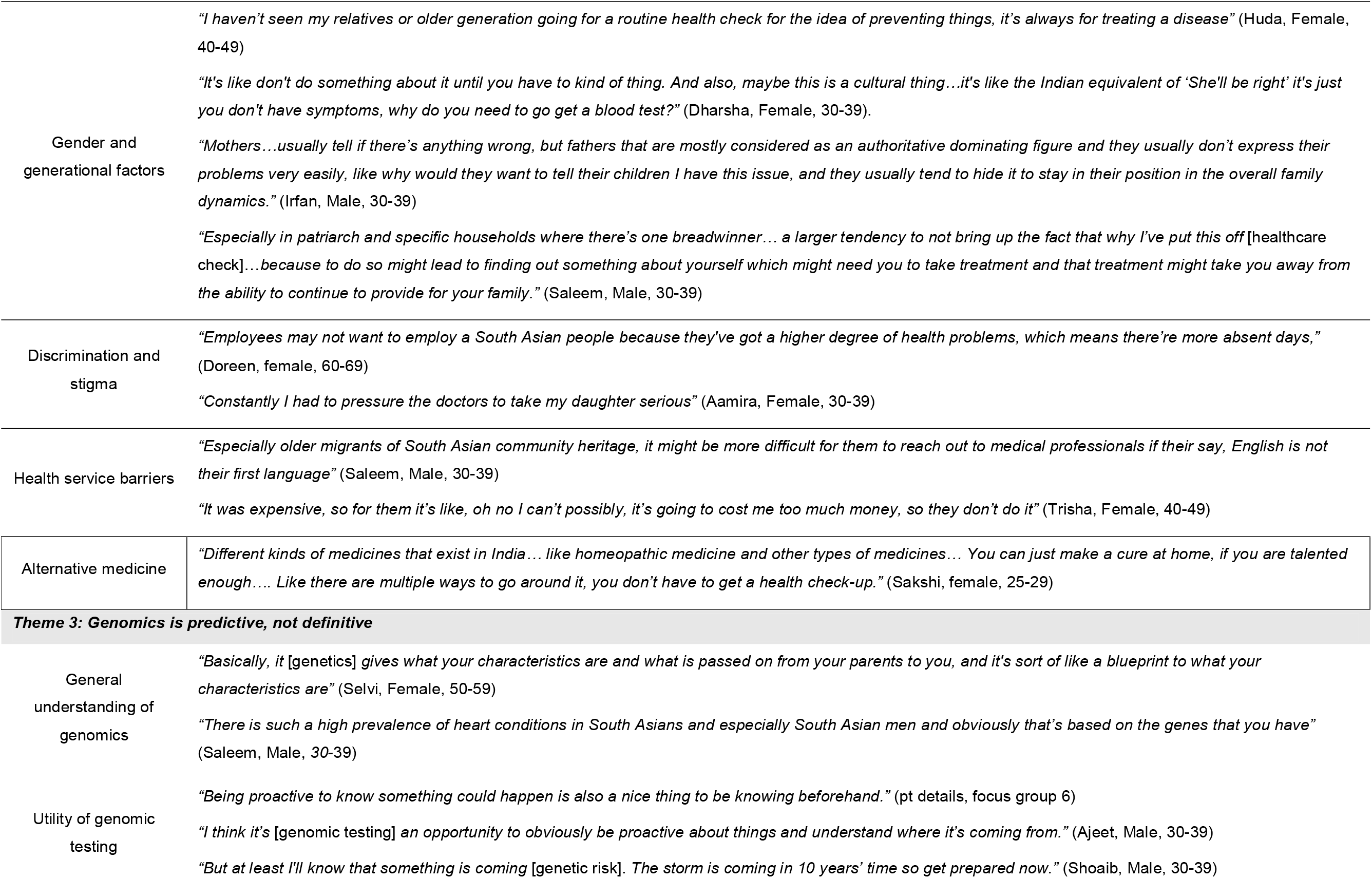
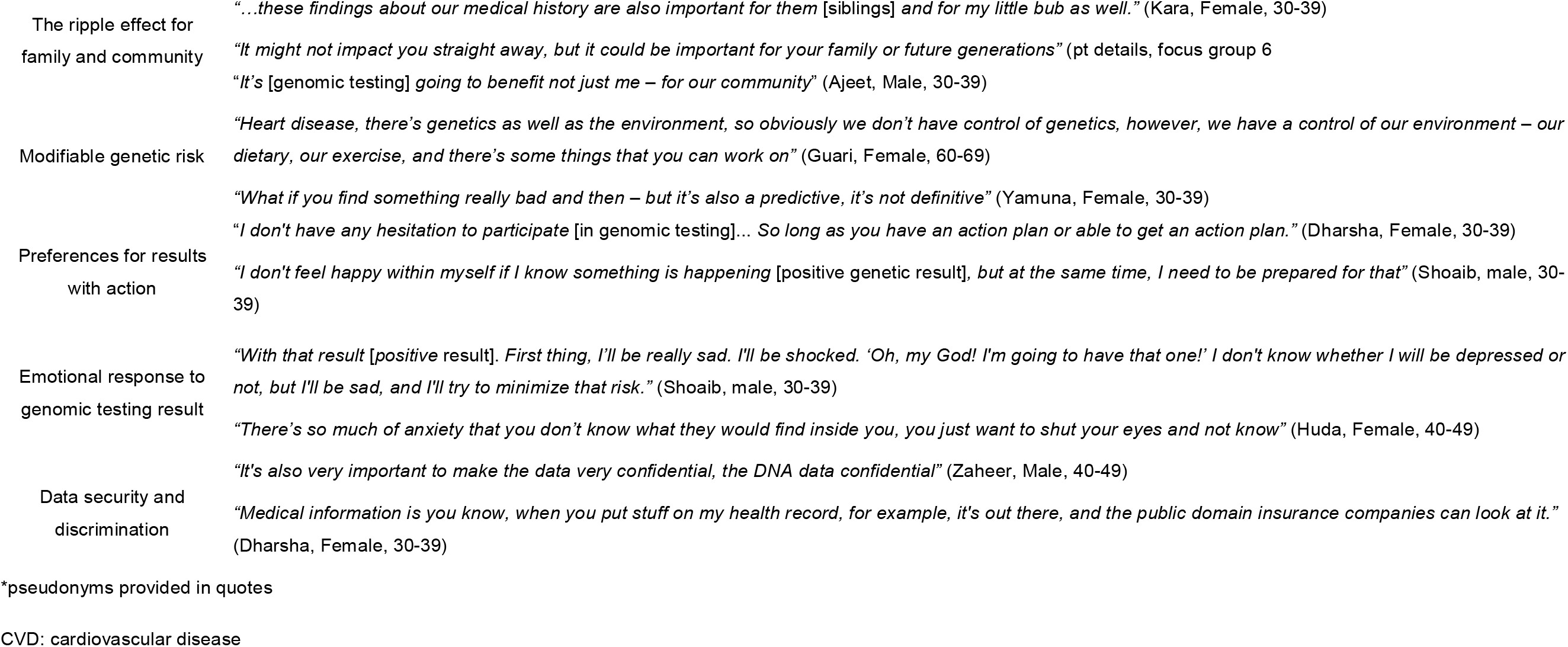
Representative quotes for study themes*.

| Topic | Representative quote |
| --- | --- |
| **Theme 1: Common thread of CVD** |  |
| Familial identity | "I don't remember anyone [in my family] who died for reason other than a heart disease" (Irfan, Male, 30-39)<br>"Heart disease is actually really prevalent in my family. My uncle had a triple bypass at 50, my grandpa died in his 50s from heart disease, like it's a huge issue." (Trisha, Female, 30-39). |
| Normalisation of risk | "I also have family history of heart disease and diabetes and all those things, the normal South Asian issues" (Aamira, Female, 30-39)<br>"We know that the prevalence of chronic disease has been dramatically increased in South Asian countries, particularly in our countries" (Zaheer, Male, 40-49) |
| CVD related worry | "This fear, disease-related fear, that tomorrow it [heart disease] might...get the better of him" (Sonu, female, 60-69)<br>"The fear is very real. Like if you – even if you don't have a history [of CVD], the fear itself is real" (Huda, female, 40-49) |
| Fatalistic beliefs | "I'm sure I will die from heart failures or some heart disease." (Doreen, female, 60-69)<br>"I think also that narrative that my dad has around, 'Oh, well, if it's going to happen, it'll happen'." (Kara, Female, 30-39) |
| Stigma | "The community and people are very uncomfortable to talk about it [heart disease risk]." (Farida, Female, 50-59)<br>"Especially in our culture there may be sort of a stigma behind it [speaking about heart disease] and judgement" (Heneesha, Female, 25-29) |
| **Theme 2: Empowering healthcare** |  |
| Modifiable risk and culture | "...a lot of it is probably our eating habits our health, you know, alcohol intake smoking type. The thing is, we've got all of this knowledge, but a lot of people are doing despite it" (Doreen, female, 60-69)<br>"It's also just general lack of physical activity in most South Asian societies..." (Saleem, Male, 30-39)<br>"Following [male relative's] heart surgery, he has made a lot of changes to his diet, which I think has been hard for him within his siblings and his own mum because the sort of meals and the different times that they eat, and they eat a lot of salt in their meals." (Kara, Female, 30-39) |
| Healthcare engagement | "Because of my family history, I got my blood checked and I have high pressure... makes me aware that I have a higher risk... I'm getting to know more about what I can do to prevent any cardiovascular disease in the future" (Zaheer, Male, 40-49)<br>"Attitude of the Indian population that's sort of thinking about [not] preventing but only do something when it's absolutely necessary to." (Lalitha, Female, 60-69) |
| Gender and generational factors | *"I haven't seen my relatives or older generation going for a routine health check for the idea of preventing things, it's always for treating a disease" (Huda, Female, 40-49)* |
|  | *"It's like don't do something about it until you have to kind of thing. And also, maybe this is a cultural thing...it's like the Indian equivalent of 'She'll be right' it's just you don't have symptoms, why do you need to go get a blood test?" (Dharsha, Female, 30-39).* |
|  | *"Mothers...usually tell if there's anything wrong, but fathers that are mostly considered as an authoritative dominating figure and they usually don't express their problems very easily, like why would they want to tell their children I have this issue, and they usually tend to hide it to stay in their position in the overall family dynamics." (Irfan, Male, 30-39)* |
|  | *"Especially in patriarch and specific households where there's one breadwinner... a larger tendency to not bring up the fact that why I've put this off [healthcare check]...because to do so might lead to finding out something about yourself which might need you to take treatment and that treatment might take you away from the ability to continue to provide for your family." (Saleem, Male, 30-39)* |
| Discrimination and stigma | *"Employees may not want to employ a South Asian people because they've got a higher degree of health problems, which means there're more absent days," (Doreen, female, 60-69)* |
|  | *"Constantly I had to pressure the doctors to take my daughter serious" (Aamira, Female, 30-39)* |
| Health service barriers | *"Especially older migrants of South Asian community heritage, it might be more difficult for them to reach out to medical professionals if their say, English is not their first language" (Saleem, Male, 30-39)* |
|  | *"It was expensive, so for them it's like, oh no I can't possibly, it's going to cost me too much money, so they don't do it" (Trisha, Female, 40-49)* |
| Alternative medicine | *"Different kinds of medicines that exist in India... like homeopathic medicine and other types of medicines... You can just make a cure at home, if you are talented enough.... Like there are multiple ways to go around it, you don't have to get a health check-up." (Sakshi, female, 25-29)* |
| **Theme 3: Genomics is predictive, not definitive** |  |
| General understanding of genomics | *"Basically, it [genetics] gives what your characteristics are and what is passed on from your parents to you, and it's sort of like a blueprint to what your characteristics are" (Selvi, Female, 50-59)* |
|  | *"There is such a high prevalence of heart conditions in South Asians and especially South Asian men and obviously that's based on the genes that you have" (Saleem, Male, 30-39)* |
| Utility of genomic testing | *"Being proactive to know something could happen is also a nice thing to be knowing beforehand." (pt details, focus group 6)* |
|  | *"I think it's [genomic testing] an opportunity to obviously be proactive about things and understand where it's coming from." (Ajeet, Male, 30-39)* |
|  | *"But at least I'll know that something is coming [genetic risk]. The storm is coming in 10 years' time so get prepared now." (Shoaib, Male, 30-39)* |
| The ripple effect for family and community | *"...these findings about our medical history are also important for them [siblings] and for my little bub as well."* (Kara, Female, 30-39)<br>*"It might not impact you straight away, but it could be important for your family or future generations"* (pt details, focus group 6)<br>*"It's [genomic testing] going to benefit not just me – for our community"* (Ajeet, Male, 30-39) |
| Modifiable genetic risk | *"Heart disease, there's genetics as well as the environment, so obviously we don't have control of genetics, however, we have a control of our environment – our dietary, our exercise, and there's some things that you can work on"* (Guari, Female, 60-69)<br>*"What if you find something really bad and then – but it's also a predictive, it's not definitive"* (Yamuna, Female, 30-39) |
| Preferences for results with action | *"I don't have any hesitation to participate [in genomic testing]... So long as you have an action plan or able to get an action plan."* (Dharsha, Female, 30-39)<br>*"I don't feel happy within myself if I know something is happening [positive genetic result], but at the same time, I need to be prepared for that"* (Shoaib, male, 30-39) |
| Emotional response to genomic testing result | *"With that result [positive result]. First thing, I'll be really sad. I'll be shocked. 'Oh, my God! I'm going to have that one!' I don't know whether I will be depressed or not, but I'll be sad, and I'll try to minimize that risk."* (Shoaib, male, 30-39)<br>*"There's so much of anxiety that you don't know what they would find inside you, you just want to shut your eyes and not know"* (Huda, Female, 40-49) |
| Data security and discrimination | *"It's also very important to make the data very confidential, the DNA data confidential"* (Zaheer, Male, 40-49)<br>*"Medical information is you know, when you put stuff on my health record, for example, it's out there, and the public domain insurance companies can look at it."* (Dharsha, Female, 30-39) |

\*pseudonyms provided in quotes
CVD: cardiovascular disease

**Figure 1.**
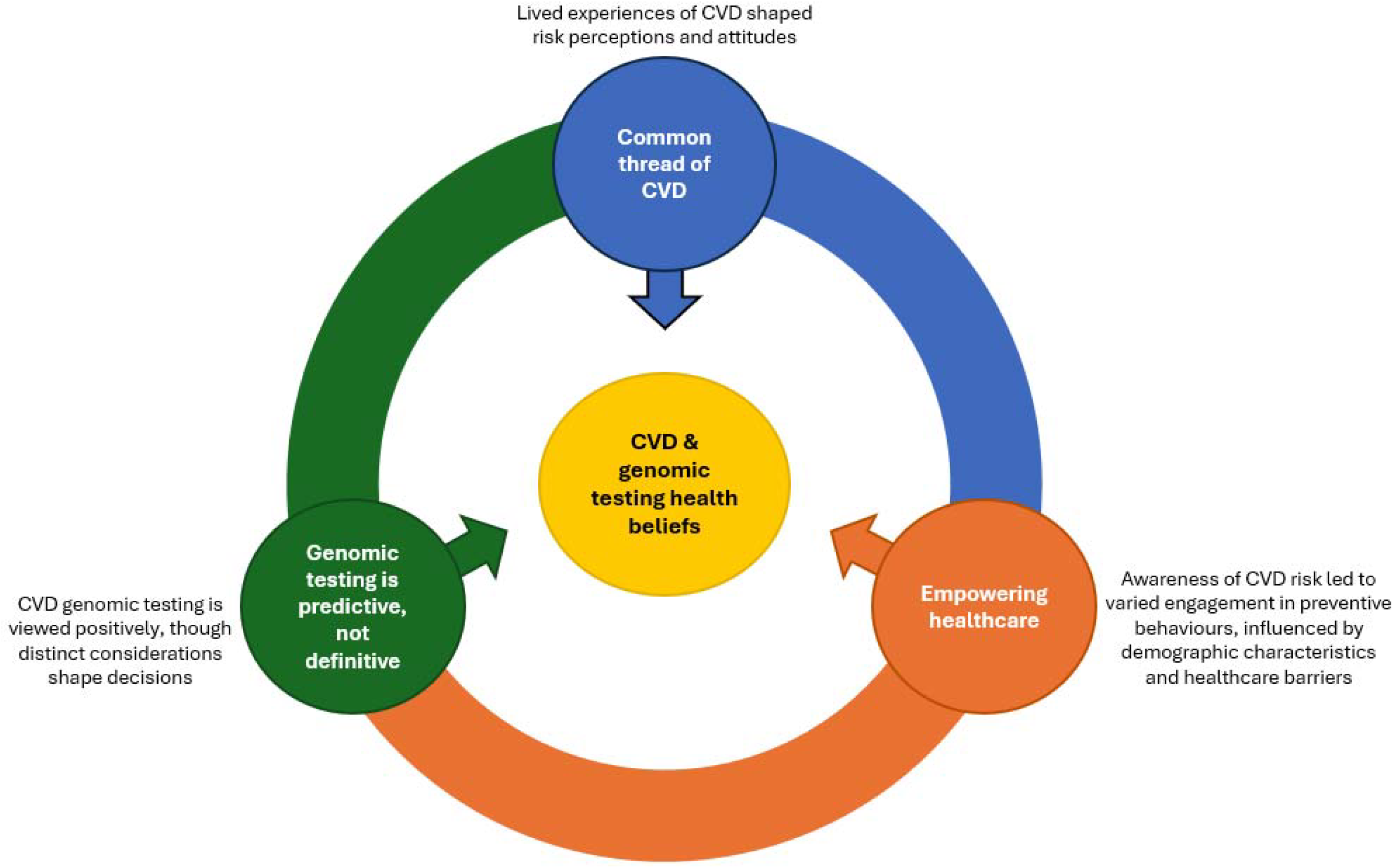
South Asian Australians’ perceptions towards CVD risk and genomic testing.

### Theme 1: Common thread of CVD

Theme 1 captures how participants’ experience with CVD shaped their perceptions of risk and disease severity. All participants had lived experience of CVD, either personally or through family members, which strongly shaped their related health beliefs. Such lived experience led many participants to describe CVD as a core aspect of their familial identity and that prevalence of risk factors such as hypertension and high cholesterol were expected with advancing age. Despite the high prevalence of CVD among participants, several noted challenges in obtaining detailed family history due to lack of open communication within families, information being lost following migration, and changes in family communication post-migration. Furthermore, some participants noted that older generations and males often did not discuss health related information, further hindering access to medical information.

Significant disease-related worry was frequently noted by most participants, with fear expressed for themselves and their family members. For some, their experiences had contributed to fatalistic beliefs regarding their susceptibility to CVD and that of their community. Although participants were willing to share their lived experience and views towards CVD within the study context, many highlighted that stigma from medical professionals and the wider community often hindered such discussions more widely.

### Theme 2: Empowering Healthcare

Theme 2 captures South Asians awareness of their increased CVD risk compared to other populations, the multifactorial nature of CVD and varied adoption of preventative measures.

Participants were largely aware that South Asians were at increased risk of CVD due to non-genetic risk factors (e.g., dietary patterns, smoking and alcohol consumption). Stress was commonly raised as an additional risk factor among migrants to Australia, attributed to adjusting to new societal and employment cultures, living away from family, and facing financial constraints. A few participants also noted that South Asians often participated in limited physical activity, particularly among women and older generations. While participants largely recognised that such lifestyle factors were modifiable, several challenges were identified to affect meaningful change. Specifically, dietary patterns and engagement in physical activity were frequently driven by cultural norms, and thus, recognised as difficult to modify.

Almost all participants understood the benefit of regular health checks and preventative behaviour. Some participants reported that awareness of their increased risk of CVD due to their South Asian background and family history motivated them to undergo regular health checks. These participants tended to value actionable health information and preventative practice. Comparatively, others preferred to delay health checks until a treatable issue arose, rather than engaging in preventative care. Younger generations and women more frequently expressed interest in adopting preventative behaviours. In contrast, males were often described as reluctant to engage in these practices, often avoiding health-related discussions and feeling the need to uphold their patriarchal role as a provider and not burden their family. Older generations were also noted to be less likely to engage in preventative behaviours, often seeking health checks only for disease treatment rather than prevention. Barriers to engaging in regular healthcare checks were commonly noted, including concerns about discrimination, time commitment, financial pressures and language barriers. Participants frequently cited challenges of navigating health systems in Australia, particularly when such systems fail to acknowledge or accommodate their cultural beliefs. Furthermore, cultural differences in health care approaches were also noted, with some individuals considering non-Western medicine and spirituality as an important component of their health care.

### Theme 3: Genomics is predictive, not definitive

Theme 3 reflects participants general understanding of the genomic contribution to CVD. Participants generally held positive views towards CVD genomic testing, recognising its potential to provide tailored risk management information for themselves, their family, and broader community. However, several participants raised concerns related to data security, insurance, and the potential cultural implications of genomic information.

At the start of focus groups, most participants demonstrated a general understanding of genomic concepts, including inheritance and its contribution to personal characteristics. Participants also acknowledge differences in CVD risk among South Asian communities were likely influenced by genetics. Participants were generally interested in undergoing genomic testing for CVD to better understand their risk and implement proactive preventative measures. Participants also recognised that the benefits of such information could extend to their family, and in some cases the wider South Asian communities. However, in select cases, participants questioned the benefits of genomic testing and research, as noted by Ajay, (male 30-39 years) at the start of the focus group: *“I can’t change my genetics, so what’s the point*”. The presentations about CVD genomics delivered during focus groups helped correct misconceptions about the utility of genomic testing and lead to more positive attitudes towards receiving genomic risk information. The information provided aligned with their pre-existing knowledge that CVD risk is influenced by both genetic and non-genetic risk factors and demonstrated the value of genomic testing. Moreover, participants noted that understanding the genomic contribution to CVD helped explain the heightened risk observed among South Asians, particularly in relation to their lived experience.

When reflecting on the potential types of genomic test results, most participants expressed a preference for receiving actionable findings only, where interventions could be implemented to mitigate the identified risks. Select participants reported they would prefer not to receive results without risk mitigative strategies, fearing that such findings would cause unnecessary fear about uncontrollable risk. Possible emotional distress from high-risk results were noted. However, such anticipated responses did not deter participants from being interested in CVD genomic testing. Participants widely recognised that genomic testing involves distinct familial considerations, unique to other clinical tests, due to its potential for familial implications. Concerns about data security, confidentially, and the potential discrimination (e.g. insurance and employer) were commonly raised. Furthermore, stigma surrounding genetic conditions was frequently noted, with participants expressing concerns about how genetic information could impact community standing, marriage prospects, and social relationships.

## Discussion

This is the first Australian study to explore the health beliefs of South Asians regarding CVD and genomic testing. South Asian Australians are recognised as an important demographic for CVD risk management (3), thus, it is essential that healthcare providers understand the unique cultural factors that influence health beliefs and healthcare engagement. Among this cohort, participants had a strong lived experience with CVD risk, which contributed to significant disease-related worry, and in some cases fatalistic beliefs. However, in line with previous research, many participants had limited details about their family history due to the lack of open discourse regarding health topics and the loss of information due to migration (24). Participants CVD health beliefs were strongly influenced by cultural norms, acculturation, and generational conflict. Such health beliefs lead to varied healthcare engagement and preventative measures. However, consistent with previous findings, significant barriers to engaging in regular health checks remain, such as fear of discrimination (25). While participants initially held some misconceptions about the utility of genomic testing, the brief educational presentations delivered during focus groups helped clarify the purposes of testing and shift attitudes.

Participants’ health beliefs were strongly informed by their lived experience of CVD. Although participants’ familial experiences of CVD heightened their risk awareness, for some it engendered fatalistic beliefs about their preordained susceptibility to CVD. Similar fatalistic attitudes have been observed among South Asian cancer and coronary heart disease cohorts, where those with fatalistic beliefs avoid engaging in preventive measures (26-28). Specifically in cancer settings, fatalism is associated with reduced uptake of cancer screening, delayed presentation to healthcare providers, and suboptimal treatment adherence (29). To address such fatalistic beliefs, future interventions addressing CVD risk among South Asian Australians should incorporate culturally informed educational strategies to increase uptake of preventative health behaviours (30, 31).

The elevated CVD risk experienced by South Asian Australians is intricately linked to their health beliefs, which, in turn, shape health behaviours. Consistent with previous literature, factors including dietary patterns, lack of physical activity, and heightened stress were commonly perceived to contribute to increased CVD susceptibility among South Asians (25, 28). Such factors are influenced by differing social priorities inherent among immigrant South Asians communities and cultural norms, which can conflict with Western concepts of health and models of behaviour change (32). Several culturally adapted lifestyle interventions have targeted CVD risk factors (e.g., blood pressure, diet, physical activity) among South Asians (33, 34). While similar interventions have been successful in the general populations, the impact of such programs has been modest within South Asian communities. Thus, highlighting the need for further research to identify how tailored lifestyle programs can most effectively reduce CVD risk among South Asian populations. Notably, community-promoted programs and the involvement of bilingual healthcare professionals of South Asian ancestry have been shown to enhance meaningful engagement (35). Given the importance of cultural norms and social values among South Asians, contemporary health interventions that focus on the individual and their self-efficacy (e.g., health belief model) may not be applicable to this population group. Rather initiatives that receive community endorsement, engage stakeholders, and prioritise the involvement of South Asian healthcare providers may be more effective in addressing the community’s needs (36). Future efforts should focus on developing frameworks to better understand the factors shaping South Asians health beliefs and their influence on the initiation and sustainment of health behaviours.

Participants general understanding of the genetic concepts and contribution to CVD risk, along with their positive attitudes toward genomic testing, align with broader findings in the literature on ancestrally diverse groups (31). As reported in other population groups, interest in genomic testing was driven by South Asians perceptions of its clinical and personal utility (37, 38). Furthermore, cultural factors were noted to have an important role in shaping attitudes toward genomic research within South Asian communities, such as the potential impact of results on community standing, marriage prospects, and social relations. Stigma secondary to genetic information remains a significant barrier to genomic testing among South Asian communities (38-41). As documented in the cancer setting, such apprehension can negatively impact communication regarding genetic health conditions within families and among South Asian communities (24, 42). Additionally, although potential emotional distress from high-risk results was acknowledged, research consistently indicates no evidence of long-term distress following the receipt of such results (43).

## Limitations

The findings of this study should be interpreted within the context of its limitations. Firstly, focus groups were publicised and conducted in English, potentially limiting participation from individuals with diverse linguistic backgrounds and hindering participants ability to fully express their views. While no interpreters were requested, some participants unexpectedly needed translational assistance and were unable to partake in the focus group. Additionally, there was limited geographical and gender diversity in the cohort with most participants identifying as female and of Indian ancestry. However, the latter is expected given that India is the most commonly reported South Asian country of birth in Australia (44). Moreover, the study cohort was characterised by a high level of educational attainment.

While such education levels may have influenced participants’ responses, it is consistent with the demographic profile of this population, which predominantly comprises highly skilled migrants residing in Australia (45). Key strengths of the study include extensive community engagement to ensure diverse views were captured where possible, inclusion of South Asian and community leaders in the research team to enhance participation and ensure a safe, respectable environment to sharing cultural nuances and experiences. Overall, our findings identify the unique cultural factors that drive engagement with CVD risk management. Findings can be used to inform future research and development of tailored strategies to improve CVD outcomes for South Asian Australians.

## Conclusions

South Asians represent a crucial demographic for CVD risk management due to the high prevalence of CVD within this group and the significant representation of this population group in Australia. This study highlights the widespread occurrence of CVD among South Asian Australians and demonstrates how lived experiences and cultural norms shape perceptions towards CVD and genomic testing, as well as engagement in preventive behaviours. Future prevention strategies should account for South Asian Australians’ health beliefs related to CVD and genomic testing.

## Data Availability

The de-identified data we analysed are not publicly available, but requests to the corresponding author for the data will be considered on a case-by-case basis.

## Acknowledgements

We would like to acknowledge members of the SAGHA Community Advisory Group for their advice and assistance in the design phase of this pilot project. Thank you to members of the Human Studies Unit, Institute for Molecular Bioscience University of Queensland, Anjali Henders, Laura Ziser and Madhura Bhadravathi Lokeshappa for the development and use of the Human Studies Research Portal database for this project. Thank you to Focus Group participants, community members of the South Asian community in Queensland, Australia for their contribution to this work.

