## Supplementary material 1 for "“*Being proactive*”: A qualitative study of South Asian Australians perspectives of cardiovascular disease and genomic testing"

The University of Queensland
Australia

CREATE CHANGE

### Focus Group:
### The South Asian Genes and Health in Australia (SAGHA) Study

### Acknowledgement of Country

The University of Queensland (UQ) acknowledges the Traditional Owners and their custodianship of the lands on which we meet.

We pay our respects to their Ancestors and their descendants, who continue cultural and spiritual connections to Country.

We recognise their valuable contributions to Australian and global society.

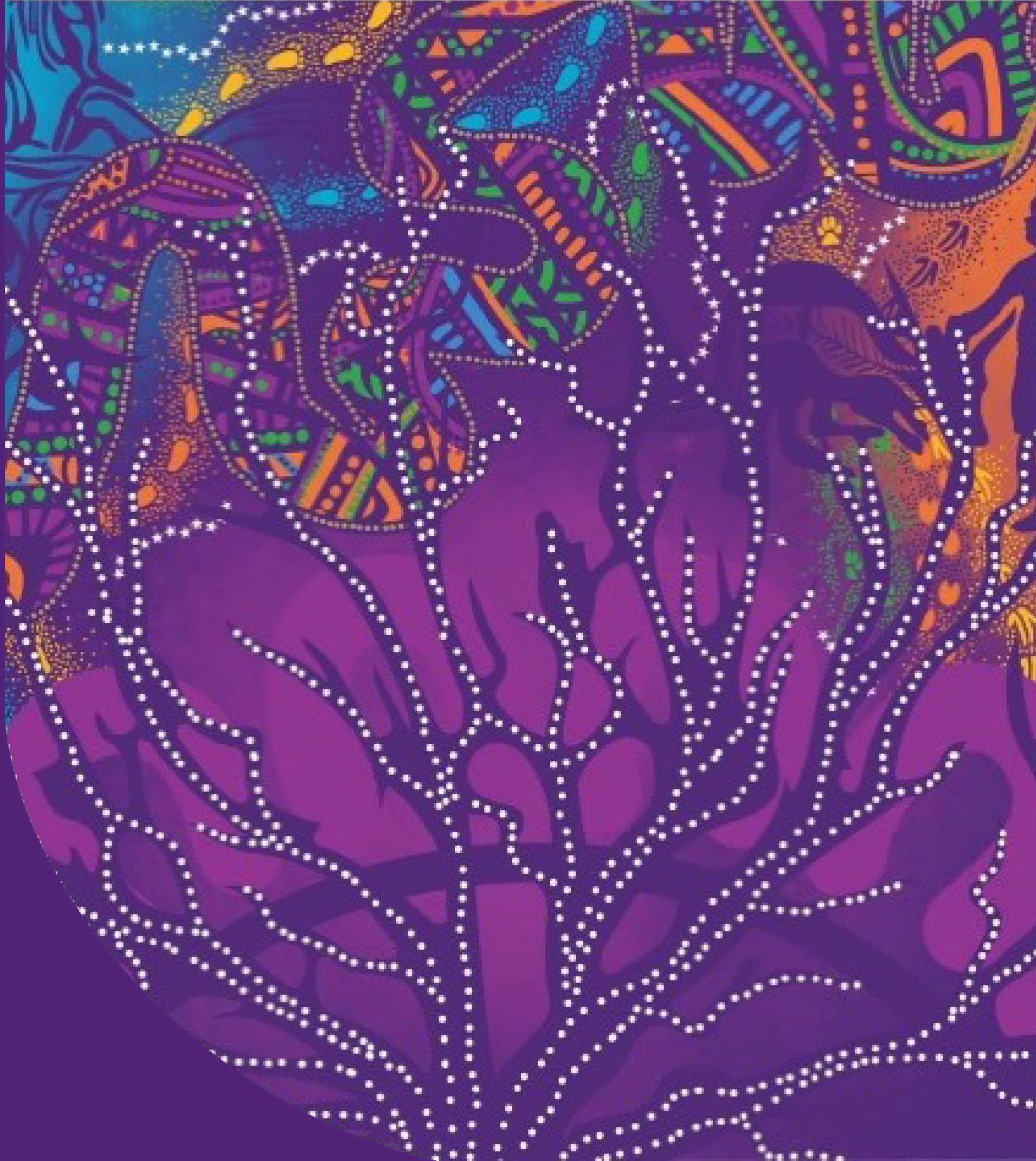

The University Of Queensland
Australia

### OUTLINE

01

**Introductions**

02

Genetics presentation

03

Genetics and heart disease

04

Conclusion

### Introduction: Who are we?

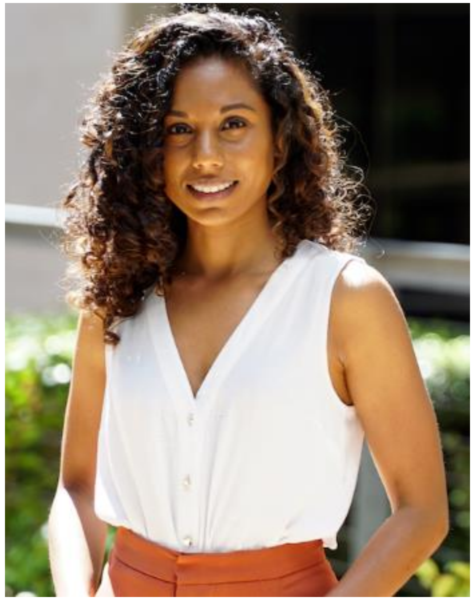

Sonia Shah
Researcher, UQ

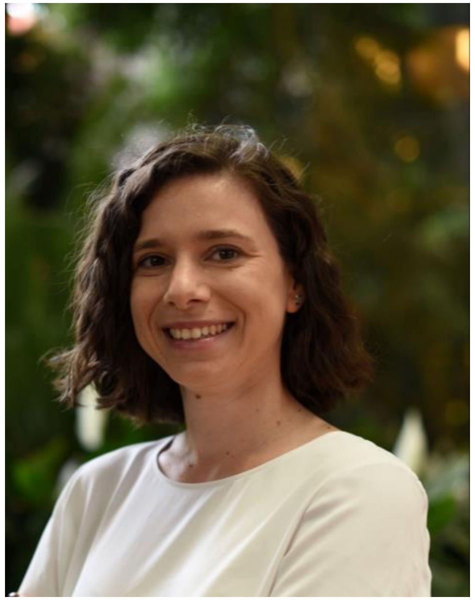

Tatiane Yanes
Genetic Counsellor
Researcher, UQ

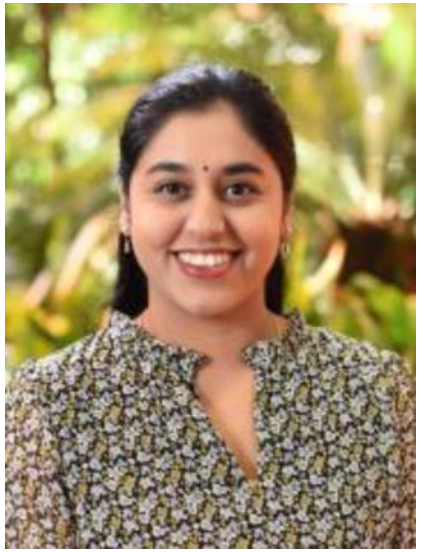

Vaish Nathan
Genetic Counsellor
Researcher, UQ

### Introduction: Purpose of the study

- Heart disease is the leading cause of death in Australia
- High impact for South Asian population

- Genetics can impact risk for heart disease

- Limited inclusion of South Asian people in health and genetics research

- How can we best work with the South Asian community to improve participation in genetics research?

### Introduction: Housekeeping

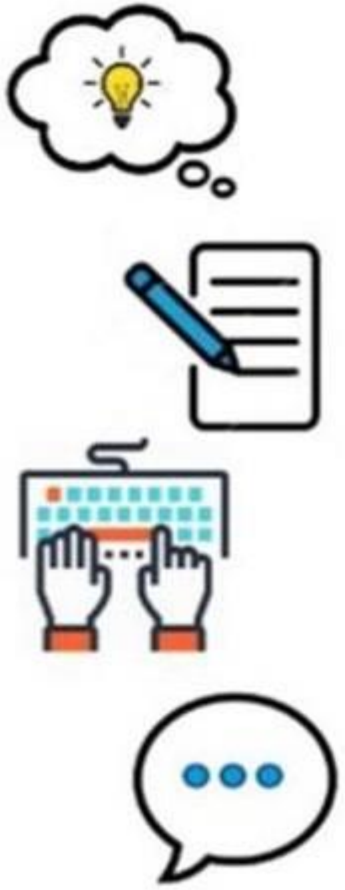

**Respect opinions**

**Recording**

**Open Discussion**

**Private and confidential**

### Introduction: Participants

1. **Can you please tell me about yourself?**
   - What sort of work do you do?
   - Tell me about your family?
   - What is your family ancestry?

2. **Have you, or anyone you know has ever been part of any health research?**

### OUTLINE

01

Introductions

02

**Genetics presentation**

03

Genetics and heart disease

04

Conclusion

### Genetics

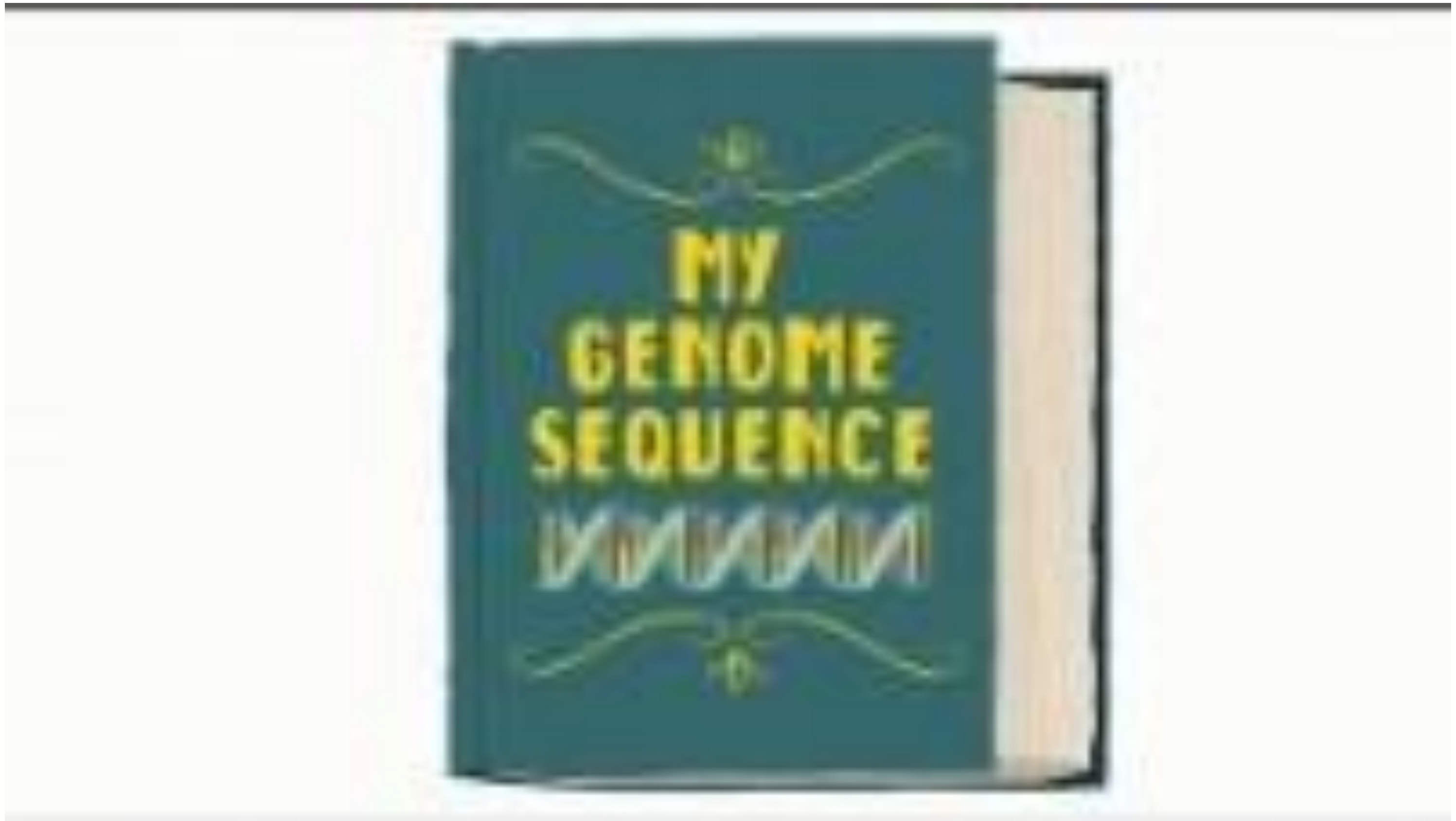

### Question

1. What are your initial thoughts after hearing this presentation?

2. What are your thoughts about participating in genetic research?

3. How do following topics influence your decision to be part of genomic research?

### OUTLINE

**01**

Introductions

**02**

Genetics presentation

**03**

**Genetics and heart disease**

**04**

Conclusion

### Polygenic risk

- Poly = many
- Genic = genetic
- Accumulation of multiple genetic "hot spots“
- Can estimate genetic risk

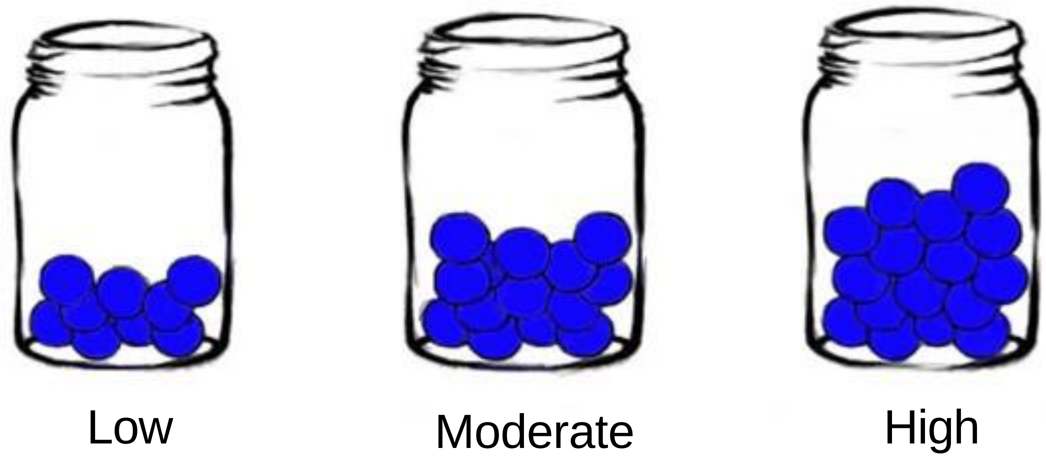

### Polygenic risk

- Poly = many
- Genic = genetic
- Accumulation of multiple genetic "hot spots“
- Can estimate genetic risk but…
  - Does not account for non-genetic factors
  - Can combine genetic and non-genetic risk factors

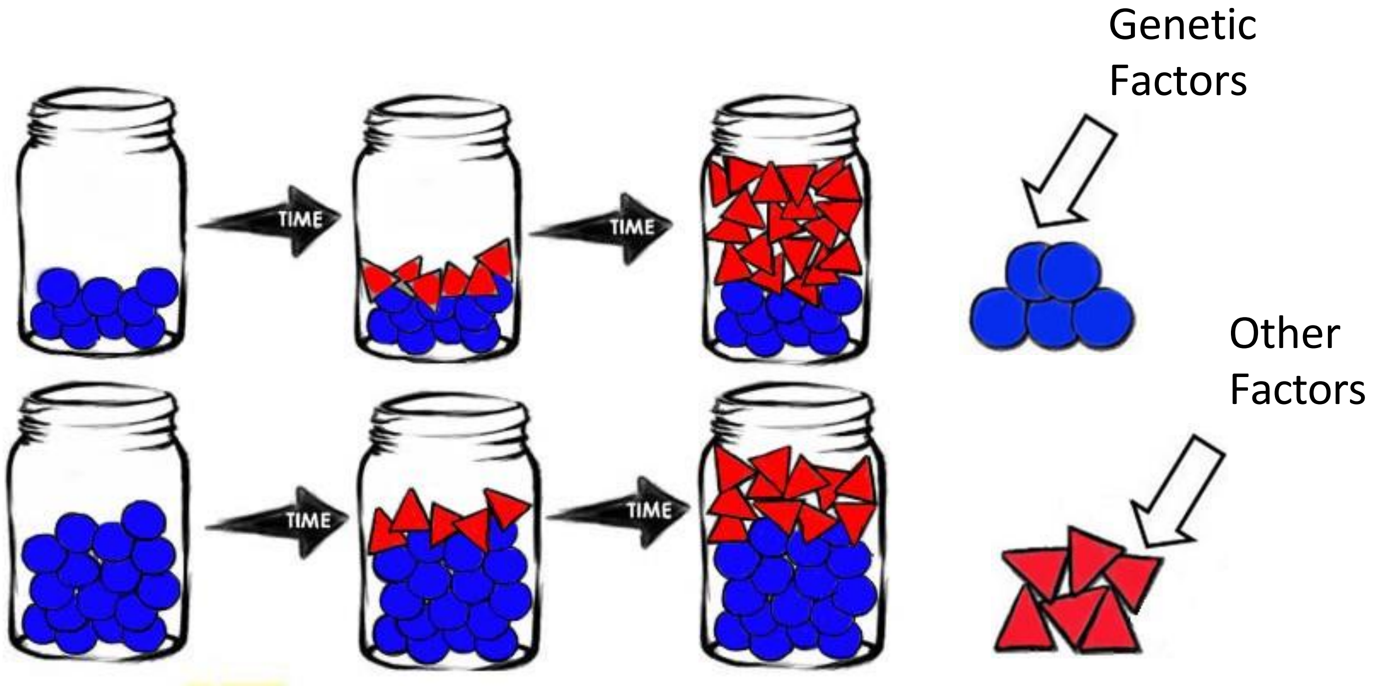

### Importance of ancestry

- South Asians make up around 23% of the world's population but...

- Only 1.3% of genetic study participants are South Asian
- Around 86% of genetic databases are based on European people

- More accurate polygenic risk for people of European ancestry
- Impacts use in healthcare

- Similar lack of diverse data across healthcare

### Question

1. How did those slides fit with your understanding about the risks for heart disease?

2. What do you think would help support South Asian community be more involved in genetic research for heart disease?

### Question

1. What would it mean to you to get a genetic test result about your risk of developing heart disease?

2. If you were invited to participate in a genetic study on heart disease, what you need to know before making a decision?

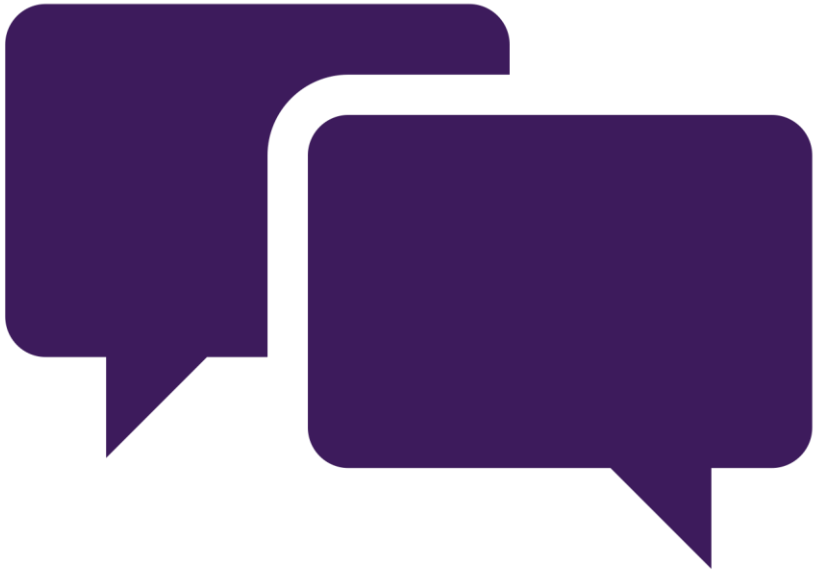

### OUTLINE

**01**

Introductions

**02**

Genetics presentation

**03**

Genetics and heart disease

**04**

**Conclusion**

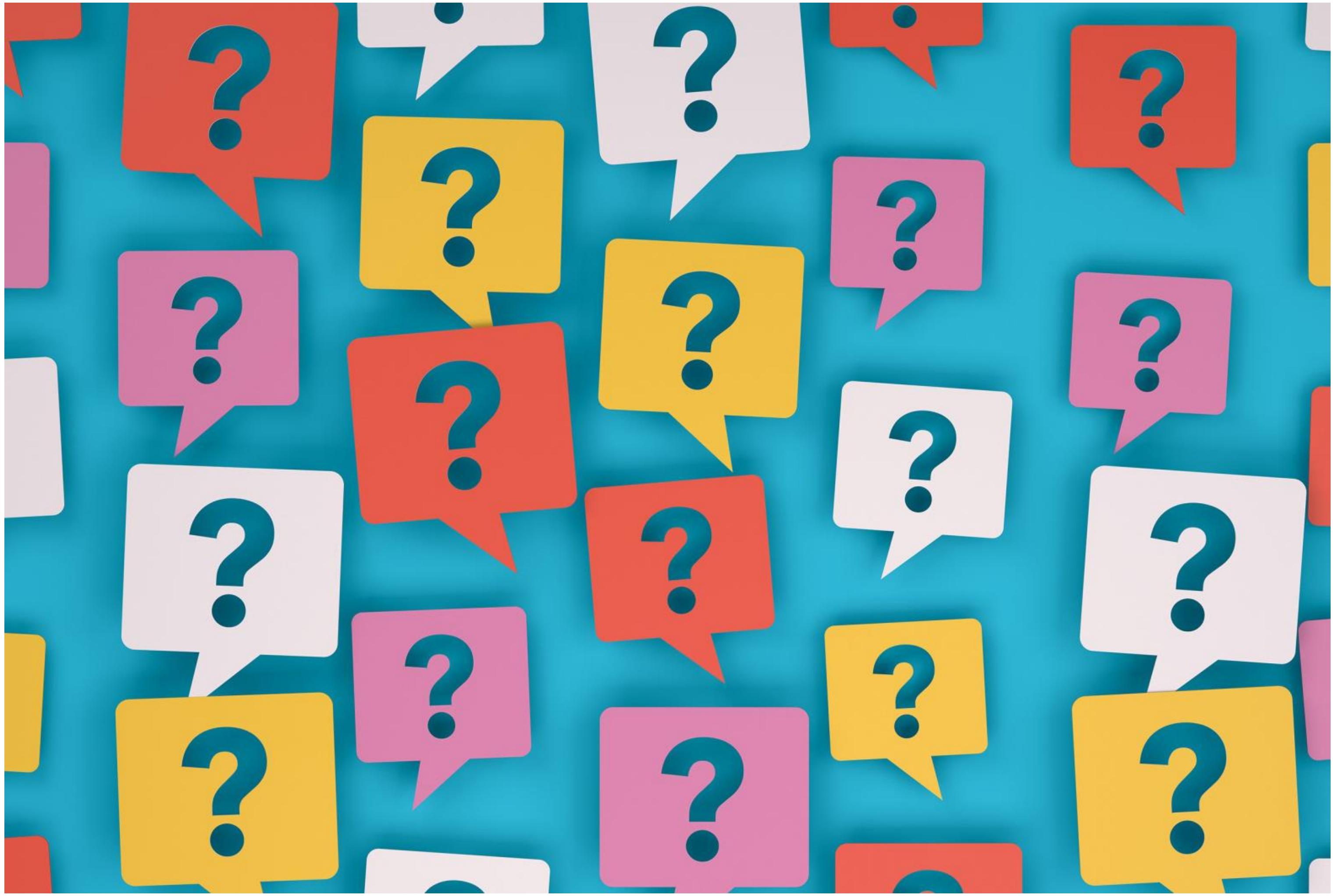

Questions?

### Thank you

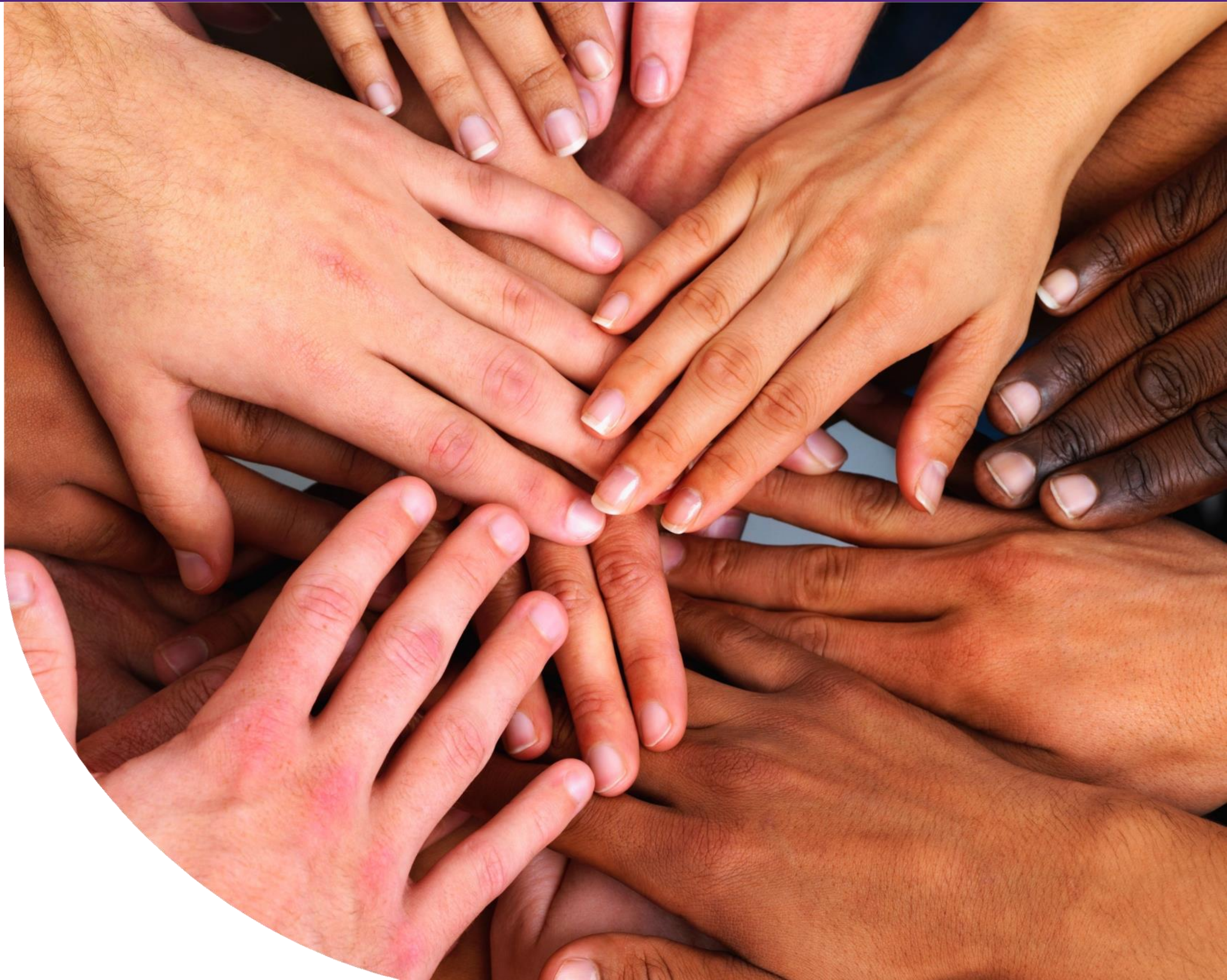
