## Supplementary material 2 for "“*Being proactive*”: A qualitative study of South Asian Australians perspectives of cardiovascular disease and genomic testing"

**Semi-structured interview Guide**

**Introduction Script**

- Thank participant for agreeing to take part in this study.
- Acknowledgement of Country
- Outline
- Briefly introduce the researcher and brief professional background.
- Introduce the study and briefly discuss its purpose of focus groups
  - Heart disease
  - South Asians
  - Genetics impact risk
  - Limited inclusion of other ethnicities
  - Engaging community
- Housekeeping
  - Reassure participants that they are free to talk about any aspect of their experience or attitudes. There are no right or wrong or even typical answers to any of the questions that we will discuss. All views will be treated with equal relevance to the research.
  - Remind participants that, with their permission, the session/interview will be recorded.
  - Reassure confidentiality and the participant’s right to leave the focus group or stop the interview at any time.
  - Clarify that the focus group will take approximately 1.5 – 2 hours and interview will take approximately 30 minutes.
  - Ask whether participants have questions before commencing.

Focus group date:

Location:

Number of participants:

| **Topic** | **Question** | **Prompt** |
| --- | --- | --- |
| ***Introduction presentation*** | *Slides:* researcher introduction, purpose of the study, role of facilitator.  Confirm participants are happy to continue with focus group  Ask if they have any additional questions | |
| **Introductions/Ice breaker** | To start, can you please tell me about yourself? | What sort of work do you do?  What is your ancestry?  Tell me about your family? |
| **Current engagement with health research** | Can you tell me if yourself, or anyone you know has ever been part of any health research? | *If yes:*  What prompted them to be part of the study?  What was the experience like?  What type of research was it? E.g. did it involve genetics?  *If no:*  What are some of the reasons why you think you have not participated in research studies? |
| **Current thoughts/views about genetics** | What do you know about genetics? | What comes to mind when you hear genetics? |
| ***Genetic presentation*** | *Slides:* background into DNA/Genes, role of DNA in health | |
| **Decision-making for genetic research, barriers and facilitators** | What are your thoughts about participating in genetic research?  *Use whiteboard to note comments* | What are the potential benefits (For yourself? For your family? For society?)?  What are the potential risks or challenges (For yourself? For your family? For society?)?  *After summarizing feedback, can ask about specific items of interest that may not have been mentioned.* |
|  | What might make you more inclined to participate in genetic research? | What role, if any, do the following topics influence your decision to be a part of genomic research?  Some people might consider…  Type of health condition/ prior experience with the condition?  Trust in the researcher?  Community engagement? |
|  | What are some things that might make it easier or harder to participate? | What made you participate in our SAGHA study? (if not already answered)  *How do you feel about providing sample? How best would you want to provide such a sample?* |
| ***Polygenic risk and heart disease presentation*** | *Slides:* Introduce polygenic risk, clinical risk factors for CVD, and limited databases of South Asian population |  |
| **Polygenic risk and heart disease presentation** | How did those slides fit with your understanding about the risks for heart disease?  *Use whiteboard to note comments* | What do you think about the role of genetics as a risk factor for heart disease?  Are there any other factors you think contributes to a person’s chance of having heart disease?  *After summarizing feedback, can ask about specific items of interest that may not have been mentioned, or something that fit stronger for them E.g. Diet?, Exercise? Smoking? Gender?)* |
|  | What do you think would help support South Asian community be more involved in genetic research for heart disease? | *Refer back to whiteboard for barrier comments and note feedback*  *After summarizing feedback, can ask about specific items of interest that may not have been mentioned e.g. Develop community trust? Sample collection (i.e. blood test)?*  *Accessibility, including recruitment site (i.e. GP vs university etc)?* |
| **Thoughts on genetic risk information and health behaviour** | What would it mean to you to get a genetic test result about your risk of developing heart disease?  *Use whiteboard to note comments* | What do you think are the benefits of receiving this information?  Do you have any concerns about receiving this information?  If someone has an increased genetic risk for heart disease, do you think there is anything that can be done to manage their chances of having heart disease?  *After summarizing feedback, can ask about specific items of interest that may not have been mentioned e.g Lifestyle changes?, Medication? Nothing?* |
| **Information for future study decision making** | If you were invited to participate in a genetic study on heart disease, what you need to know before making a decision?  *Use whiteboard to note comments* | *After summarizing feedback, can ask about specific items of interest that may not have been mentioned e.g Sample collection, Privacy, Consent, Understanding of genetics etc*  *Prompts:*  *What might stop you from participating?*  *What might prevent you from making the decision?*  We will be evaluating some consent forms in the future. We will invite everyone who has participated in these focus groups to review the documents. If this is not something you are interested in, please let us know. |
| **Concluding presentation** | *Slides:* Thank them for being involved in the research. Reaffirm confidentiality of the study. Provide contact information if they have any question and that we will provide a summary of the study once data collection completed. Given them Coles/Myer voucher for participation. | |
